# Ancestry Calibration of Polygenic Risk Scores Improves Risk Stratification and Effect Estimation in African American Adults

**DOI:** 10.1101/2025.06.18.25329573

**Authors:** Luciana B. Vargas, Mariah C. Meyer, Iain R. Konigsberg, Aastha Kakar, Patrick M. Carry, Polygenic Risk Methods in Diverse Populations (PRIMED) Consortium, Yun Li, Alana C. Jones, Hemant K. Tiwari, Vinodh Srinivasasainagendra, Nicole D. Armstrong, Eimear E. Kenny, Bogdan Pasaniuc, Marguerite R. Irvin, Michael H. Cho, Maggie A. Stanislawski, Sridharan Raghavan, Jonathan A. Shortt, Leslie A. Lange, Ethan M. Lange

## Abstract

Polygenic risk score (PRS) distributions vary across populations, complicating PRS risk assessment. We evaluated the impact of *post-hoc* PRS calibration according to individualized genetic ancestry estimates on PRS performance using two large multi-ethnic PRS for type 2 diabetes (T2D) (PRS_T2D_) and height (PRS_height_), in 8,841 African American (AA) individuals from the Reasons for Geographic and Racial Differences in Stroke (REGARDS) study. We calibrated each participant’s score as a function of estimated genetic similarity to the Yoruba (GSYRI) cohort in the 1000 Genomes Project. Uncalibrated PRSs were significantly skewed by GSYRI. After calibration, 33.6% of individuals in the top decile of PRS_T2D_ were reclassified and performance in the top PRS_T2D_ decile improved from an OR of 7.97 [6.31–10.13] to 10.77 [8.41– 13.91] when compared to the lowest decile. Similarly, 55.0% of individuals in the top PRS_height_ decile were reclassified with GSYRI calibration. The calibrated PRS_height_ showed higher correlation with height (from 0.24 to 0.32, *p*<10^−7^), and increased mean height in the top PRS_height_ decile (*p*=5.7×10^−5^) when compared to the uncalibrated PRS_height_. Lastly, we show that evaluating uncalibrated PRS while adjusting for GSYRI in regression models can lead to inflated and unstable effect size estimates for both the PRS and GSYRI.

## Introduction

Polygenic risk scores (PRS), which aggregate risk estimates across genetic variants associated with a trait of interest, are powerful tools for identifying individuals at increased risk for disease. Individuals with a PRS score in the extreme upper tail of the corresponding PRS distribution can have a similar risk for disease as an individual carrying a rare high-penetrant mutation^1^.

Differences in allele frequencies and linkage disequilibrium can result in shifts in the distributions of values for the same PRS both within, for genetically admixed individuals, and between populations, making it difficult to assess risk when combining different populations in the same study^2–7^. PRS calibration has been proposed as a solution to regress out the impact of ancestry on PRS. Recent methods for calibrating PRS have been proposed to address these distributional issues, often relying on standardization based on population labels, or, preferably, by regressing out genetic differences in allele frequencies (e.g., using principal components [PCs]) from these scores^8,9^. Here, we explore the implications of *post-hoc* calibration of PRS values according to individualized estimated ancestry proportions in self-identified African American (AA) individuals. We selected PRS functions for type 2 diabetes (T2D) and height, as these traits represent well-studied and common examples of complex human traits. Height is commonly used as a benchmark for genetic studies due to being a classic complex trait (i.e., influenced by thousands of genetic variants with small effect sizes) with broad availability and minimal phenotypic error across studies. Moreover, height is one of the most heritable human complex traits, with 80-90% of the variability in height estimated to be heritable^10–12^. PRS_height_ explains over 90% of SNP-based heritability for height, the highest among all available PRS to date^12^. T2D, on the other hand, is a highly prevalent condition, affecting 11.1% of the global adult population^13^, and is a major risk factor for other complex diseases. For example, T2D patients have over 50% increased risk of developing cardiovascular disease and dementia compared to those without T2D^13^. We show that *post-hoc* calibration is important when assessing PRS performance (e.g., R^2^ or AUC), when ranking individuals according to their PRS values, and when interpreting effect sizes of both estimated ancestry and PRS in relation to a phenotype.

## Methods

We leveraged genetic data from 8,841 self-identified non-Hispanic Black study participants from the Reasons for Geographic and Racial Differences in Stroke (REGARDS) cohort. REGARDS is a U.S. national prospective cohort designed to study the reasons for higher incidence of stroke and related comorbidities in African American (AA) individuals and those living in the Southeastern U.S^14^.

From the PGSCatalog^15^, we identified two recent PRS based on some of the largest multiethnic GWAS studies to date: one for type 2 diabetes (T2D) (PGS002308)^16^ and one for height (PGS002802)^12^. Both PRS_T2D_ and PRS_height_ were developed including 1.1M and 5.3M samples of different ancestral backgrounds (although >75% of primarily European ancestry), and accounted for population structure by applying different meta-analysis approaches, PRS-CSx^17^ (PRS_T2D_) and SBayesC^18^ (PRS_height_). We evaluated the performance of PRS_height_ in relation to height (inches), and PRS_T2D_ in the context of T2D status and in relation to fasting glucose levels (mg/dL) in non-diabetic individuals. Further details can be found in **Supplemental Materials**.

We characterized genetic diversity in REGARDS by estimating the fraction of an individual’s genome that comes from *K* hypothetical ancestral populations using ADMIXTURE (v. 1.3)^19,20^. Based on the prior knowledge that AA individuals have genetic admixture of primarily West African and European continental ancestry^21^, we predefined K=2 and used 1,000 Genomes reference panels (Yoruba in Ibadan, Nigeria [YRI] and Northern Europeans from Utah [CEU])^22^ to supervise ADMIXTURE estimation. Based on this method, each REGARDS individual was assigned two *K* fractions corresponding to the estimated proportion of genetic similarity to the YRI (GSYRI) and CEU reference panels (**Sup. Fig. 1**). To validate our estimates, we additionally estimated genetic similarity by calculating principal components (PCs) derived from genetic data in PLINK (v2)^23^. We assessed GSYRI both continuously and by defining bins of genetically similar individuals: >90%, 80-90%, 70-80%, 60-70%, 50-60%, and <50% (the lower deciles (0-50%) were collapsed due to small sample size, N = 246).

To remove the PRS and GSYRI correlation, we calibrated each individual’s PRS value by regressing out their GSYRI and dividing the residuals by the standard deviation of the PRS values for all participants within the same GSYRI-defined bin (to account for the possibility that variance in the PRS is also a function of ancestry proportions). This PRS calibration by genetic similarity can be represented as:

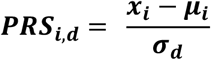

Where ***i*** is REGARDS individual ***i*; *d*** is the GSYRI decile grouping for individual ***i*; *x***_***i***_ is the observed unscaled PRS for person ***i*; *μ***_***i***_ is the predicted PRS for person ***i*** (based on regression model of *PRS ~ GSYRI*); and ***σ***_***d***_ is the estimated standard deviation of PRS scores for REGARDS individuals in GSYRI decile ***d***. To validate our approach, we compared PRS_height_ residuals after regressing out either i) GSYRI, ii) PCs 1-10, or iii) using the Polygenic Score Catalog Calculator^24^ ancestry normalization tool (*pgsc_calc*) (**Sup. Fig. 2**). The *pgsc_calc* tool performs PC projections based on the 1,000 Genomes sample space and continuously adjusts PRS values based on differences in the mean and variance of the PRS across the PC sample space^9,25^.

We compared overall model prediction and effect estimates for calibrated and uncalibrated PRS through a series of linear (height, glucose) and logistic (T2D) regression models. All models were adjusted for age and sex. T2D and glucose models were further adjusted for body mass index (BMI). For each outcome and each PRS (uncalibrated or calibrated), models were constructed including a) basic covariates, b) basic covariates and GSYRI, c) basic covariates and PRS, d) basic covariates, GSYRI and PRS, and e) basic covariates, GSYRI, PRS, and an interaction between GSYRI and PRS.

## Results and Discussion

Individuals in REGARDS that self-identified as Black have a median GSYRI of 84.35% [IQR: 75.68 - 90.63%] (**Sup. Fig. 1 and Sup. Table 1**), which agrees with other AA groups previously described^26,27^. GSYRI and the first PC are highly correlated (r = 0.9989, *p* < 10^−250^) in REGARDS data. When PRS functions were applied to REGARDS participants, we observed that both uncalibrated PRS (PRS_height_ and PRS_T2D_) were correlated with GSYRI (|r| > 0.25, *p* < 10^−16^) (**Fig. 1**). We compared PRS calibration using GSYRI, PCs 1-10, or the *pgsc_calc* ancestry-normalization tool, and the three approaches were all highly correlated (r > 0.98, *p* < 10^−16^) (**Sup. Fig. 2**). Thus, we opted to use GSYRI in our analyses, as this measure is more interpretable than PCs and has been independently shown to perform similarly to correction based on PCs^28^.

**Figure 1.**
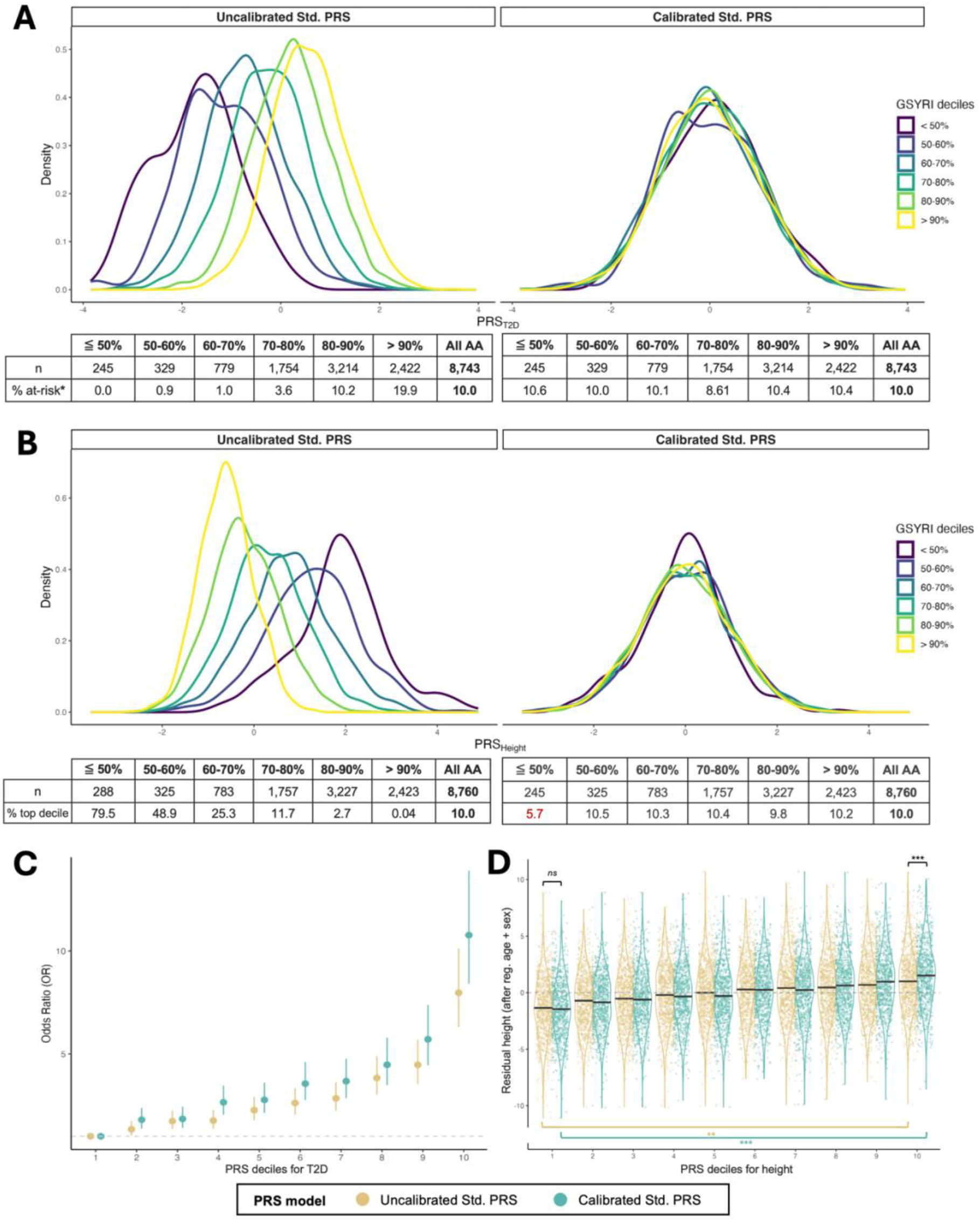
Impact of ancestry-informed calibration on PRS distribution and prediction. Distribution curves for **A)** PRS_T2D_ and **B)** PRS_height_ across African Americans (AA) individuals from the REGARDS cohort, stratified according to GSYRI, before and after PRS calibration. *At-risk was defined as individuals in the upper 10% of PRS distribution. **C)** Point estimates and confidence intervals of the odds ratio (OR) of type 2 diabetes (T2D) when comparing each decile of the PRS distribution to the lowest decile, for the uncalibrated PRS_T2D_ (yellow) and the calibrated PRS_T2D_ (blue). **D)** Violin plots showing the distribution of height (after adjusting for age and sex) across deciles of the PRS_height_ distribution, when using the uncalibrated PRS_height_ (yellow) and the calibrated PRS_height_ (blue).

It has been suggested that the reason for PRS distributional differences across populations is primarily due to differences in allele frequencies and linkage disequilibrium patters across populations^5,6,29^. An alternative explanation, however, is that real differences in genetic architecture of traits in different populations lead to the observed PRS shifts. We investigated these hypotheses by studying the association of the outcomes included in our study (height, T2D, and glucose) and GSYRI. Height and glucose levels were not associated with GSYRI (after adjusting for sex, age, [and BMI in glucose models], *p* > 0.05) while T2D was positively associated with GSYRI (after adjusting for sex, age and BMI, *β* = 1.10, *p* < 10^−07^). The latter observation has been previously reported in the literature, and it remains unclear whether the association is due to genetic differences between groups, or due to confounding environmental differences, such as social determinants of health (SDoH)^30–35^. We further adjusted T2D models for three SDoH variables (income, education, and medical care), and the association between T2D and GSYRI was attenuated (*β* = 0.70, *p =* 0.01). When stratified by sex, the association between T2D and GSYRI attenuated more strongly in males (*β* = 0.52, *p =* 0.13) than females (*β* = 0.85, *p =* 0.0034), consistent with previous reports^36^. These results suggest that, in our study, shifts in the PRS distributions associated with GSYRI are primarily due to population allele frequency differences rather than genetically driven differences in height or T2D between individuals with different GSYRI profiles.

### Ancestry Calibration Improves Risk Stratification

Based on observed values in the upper tail of the PRS distributions, the uncalibrated PRS functions either systematically under- or over-estimated risk for AA individuals with lower estimated GSYRI relative to the remaining AA individuals **(Fig. 1A-B)**. For PRS_T2D_, all AA individuals with ≦50% GSYRI were systematically excluded from the upper 10% of the uncalibrated PRS distribution “at-risk” classification (none had a PRS in the upper decile), and most (over 99%) of the individuals with ≦70% GSYRI were also excluded. The opposite shift was observed for PRS_height_, where the majority (79.5%) of individuals with ≦50% GSYRI were placed at the top decile of the uncalibrated PRS_height_ **(Fig. 1B)**, although height itself is not associated with GSYRI (*p* > 0.05).

After calibration of PRS_T2D_, 33.6% of individuals in the top decile of PRS_T2D_ distribution were reclassified. We found a higher median percentile change in PRS_T2D_ rank with calibration in the subset of individuals previously excluded from risk stratification (≦70% GSYRI) compared to the remaining individuals (>70% GSYRI) (26.7 increase versus 4.5 decrease, respectively, *p* < 0.001) (**Sup. Table 2 and Sup. Fig. 4**). Calibration increased the odds ratio (OR) of T2D from 7.97 [95% CI:6.31-10.13] when using the uncalibrated PRS_T2D_ to 10.77 [95% CI: 8.41-13.91] using the calibrated PRS_T2D_ (*p* = 0.032), when comparing dichotomized PRS (upper 10% vs. lower 10%) (**Fig. 1C)**. This reclassification also resulted in higher correlation with glucose levels (after adjusting for sex and age) in non-diabetics, from 0.083 for the uncalibrated PRS_T2D_ (*p* = 2.4 × 10^−9^) to 0.10 for the calibrated PRS_T2D_ (*p* = 2.2 × 10^−13^) (**Sup. Fig. 3A**).

After calibration of PRS_height_, 55.0% of individuals in the top decile of the uncalibrated PRS distribution were reclassified using the newly calibrated PRS rankings. Individuals ranked in the top decile of the calibrated PRS_height_ had significantly higher median height as compared to the uncalibrated PRS_height_ (median residual height percentiles: 64.3 [IQR: 38.2-84.5] and 71.7 [IQR: 46.7-88.6], *p* = 5.7 × 10^−5^) (**Fig. 1D**). Moreover, the correlation between PRS_height_ and height (after adjusting for age and sex) increased from 0.24 to 0.32 after GSYRI calibration (*p* < 10^−7^) (**Sup. Fig. 3B**). As expected, we observed that individuals with greater levels of genetic admixture had greater percentile rank changes between the PRS_height_ distributions before and after the calibration. Individuals with ≦70% GSYRI had a median 28.0 percentile decrease in PRS_height_ after calibration, compared to a median of 4.2 percentile increase in the remaining individuals (>70% GSYRI) (*p* < 0.001) (**Sup. Table 2 and Sup. Fig. 5**).

### Ancestry: Covariate Adjustment versus Calibration on Effect Estimates

It is common practice to model genetic similarity (e.g., GSYRI or PCs) as a covariate when evaluating the performance of uncalibrated PRS values to account for possible confounding between genetic similarity and the PRS. We demonstrate that, for some PRS functions, this practice alone does not remedy biased PRS effect estimates in admixed groups, such as AA individuals. We found similar overall model performance (AUC/R^2^) between prediction models including calibrated or uncalibrated PRS when also including GSYRI (Table 1 and Sup. Table 3). However, we observe biased parameter estimates for both GSYRI and the uncalibrated PRS when jointly including those in the same model. For example, GSYRI was not associated with height in models adjusted for age and sex (*β* = 0.21 ± 0.23, *p* = 0.37). However, when including the uncalibrated PRS, GSYRI became strongly associated with height (*β* = 5.69 ± 0.28, *p* = 8.4 × 10^−88^)(**Table 1**). Furthermore, the effect estimates for the *uncalibrated* PRS changed considerably depending on whether GSYRI was or was not included in the model (from *β*=0.68 ± 0.03 to *β*=1.13 ± 0.04). In contrast, when including the *calibrated* PRS in the model along with GSYRI, the effect estimate for GSYRI did not change, remaining nonsignificant, and the effect estimate for the calibrated PRS also was not impacted by the inclusion of GSYRI. A similar pattern of results was observed for PRS_T2D_ and glucose levels (**Sup. Table 3**). Of note, GSYRI was associated with T2D before inclusion of the PRS_T2D_ in the models (*β* = 1.10 ± 0.21, *p* = 7.5 × 10^−8^). However, the direction of the GSYRI effect changed when including the *uncalibrated* PRS_T2D_ in the model (from *β*=1.10 ± 0.21 to *β*=**-**2.68 ± 0.26), whereas the effect estimates for GSYRI remained similar when including the *calibrated* PRS in the model (**Table 1**). We demonstrate that, for some PRS functions, covariate adjustment for uncalibrated PRS estimates results in biased effect estimates, and that *post-hoc* calibration of PRS based on GSYRI results in more stable estimates.

**Table 1.** Change in effect sizes of PRS_height_ and PRS_T2D_ with the inclusion of estimated genetic ancestry (GSYRI) as a covariate in the models. T2D, type 2 diabetes. β, effect size of one unit change in the outcome. s.e., standard error. VIF_PRS_, variance inflation factor for the PRS in the model. R^2^, fraction of variance explained by the model. AUC, area under the receiver operating characteristic (ROC) curve. For interaction models (noted by the inclusion of the interaction term GSYRI x PRS), parameter estimates are given for each of the main effects (GSYRI or PRS) and the interaction effect (GSYRI x PRS).

|  | GSYRI |  | PRS |  | VIF<sub>PRS</sub> | R<sup>2</sup> (%) or<br>AUC |
| --- | --- | --- | --- | --- | --- | --- |
| | $\beta \pm \text{s.e.}$ | p-value | $\beta \pm \text{s.e.}$ | p-value | | |
| Height |  |  |  |  |  |  |
| ~ Age + Sex | - | - | - | - | - | 50.00 |
| + GSYRI | 0.21 ± 0.23 | 0.37 | - | - | - | 50.00 |
| + Uncalibrated PRS | - | - | 0.68 ± 0.03 | 6.97 × 10<sup>-116</sup> | 1.00 | 52.94 |
| + GSYRI + Uncalibrated PRS | 5.69 ± 0.28 | 8.44 × 10<sup>-88</sup> | 1.13 ± 0.04 | 1.72 × 10<sup>-201</sup> | 1.63 | 55.02 |
| + GSYRI + Uncalibrated PRS + (GSYRI x Uncalibrated PRS) | 1.55 ± 0.060 | 1.61 × 10<sup>-69</sup> | 0.70 ± 0.15 | 1.46 × 10<sup>-6</sup> | - | 55.07 |
| | Interaction: $\beta \pm \text{s.e.} = 0.55 \pm 0.18, p = 0.0023$ | | | | | |
| + Calibrated PRS | - | - | 0.87 ± 0.03 | 1.48 × 10<sup>-202</sup> | 1.00 | 55.04 |
| + GSYRI + Calibrated PRS | 0.22 ± 0.22 | 0.32 | 0.87 ± 0.03 | 1.48 × 10<sup>-206</sup> | 1.00 | 55.05 |
| + GSYRI + Calibrated PRS + (GSYRI x Calibrated PRS) | 0.22 ± 0.22 | 0.32 | 1.19 ± 0.19 | 3.27 × 10<sup>-10</sup> | - | 55.06 |
| | Interaction: $\beta \pm \text{s.e.} = -0.40 \pm 0.23, p = 0.083$ | | | | | |
| T2D |  |  |  |  |  |  |
| ~ Age + Sex + BMI | - | - | - | - | - | 0.65 |
| + GSYRI | 1.10 ± 0.21 | 7.50 × 10<sup>-8</sup> | - | - | - | 0.65 |
| + Uncalibrated PRS | - | - | 0.69 ± 0.03 | 2.51 × 10<sup>-121</sup> | 1.02 | 0.71 |
| + GSYRI + Uncalibrated PRS | -2.68 ± 0.26 | 6.37 × 10<sup>-25</sup> | 0.88 ± 0.04 | 8.95 × 10<sup>-137</sup> | 1.50 | 0.73 |
| + GSYRI + Uncalibrated PRS + (GSYRI x Uncalibrated PRS) | -2.57 ± 0.26 | 2.54 × 10<sup>-21</sup> | 0.68 ± 0.16 | 2.44 × 10<sup>-05</sup> | - | 0.73 |
| | Interaction: $\beta \pm \text{s.e.} = 0.24 \pm 0.19, p = 0.021$ | | | | | |
| + Calibrated PRS | - | - | 0.70 ± 0.03 | 3.16 × 10<sup>-137</sup> | 1.02 | 0.72 |
| + GSYRI + Calibrated PRS | 1.22 ± 0.21 | 1.16 × 10<sup>-8</sup> | 0.70 ± 0.03 | 9.16 × 10<sup>-138</sup> | 1.02 | 0.73 |
| + GSYRI + Calibrated PRS + (GSYRI x Calibrated PRS) | 1.11 ± 0.22 | 4.92 × 10<sup>-7</sup> | 0.36 ± 0.19 | 0.058 | - | 0.73 |
| | Interaction: $\beta \pm \text{s.e.} = 0.42 \pm 0.23, p = 0.064$ | | | | | |

The reason for unstable effect estimates in models that include uncalibrated PRS and GSYRI is not clear. We found low to moderate multicollinearity between GSYRI and the uncalibrated PRS values (variance inflation factor, VIF=[1, 1.5]), which indicates that the correlation between PRS and GSYRI (|*r*| > 0.25) does not fully explain the unreliability of the estimates observed. For height and T2D, we found no significant evidence supporting an interaction between GSYRI and PRS values in models including the calibrated PRS, but did find such evidence for modest interaction effects using the uncalibrated PRS (**Table 1**). We found no evidence of an interaction between GSYRI and PRS in models with glucose level as the outcome (**Sup. Table 3**). An alternative hypothesis to explain the unreliability of models with the uncalibrated PRS and GSYRI would be that there is a source of collider bias that is removed by directly regressing out the correlation between PRS and GSYRI (i.e., calibration) before inclusion in the regression models, although further investigation would be needed to confirm this.

**In conclusion**, this work demonstrates that *post-hoc* calibration of PRS scores using a continuous estimate of genetic similarity (e.g., GSYRI or PCs) is *needed* for more accurate interpretation and clinical utility of PRS in AAs. Although we show that overall model performance (as measured by R^2^ or AUC) when including both GSYRI and continuous PRS in prediction models is similar for both calibrated and uncalibrated PRS, there was an instability of coefficient estimates for both the PRS and GSYRI when using uncalibrated PRS. Moreover, identification of “at-risk” individuals, as would be applied in a clinical setting, differed substantially with calibration. This observation highlights the need to disentangle the impact of genetic similarity on PRS to improve interpretation of PRS as a stand-alone risk factor in genetically admixed populations. It is critical to highlight that the calibration does not *fully* address the current differences in PRS performance across the genetic ancestry continuum, and there are several other limitations that must be addressed before clinical implementation of PRS is optimized in genetically admixed individuals that were not included in the scope of this work^37^. Existing PRS calibration methods rely on broadly regressing out measures of global ancestry (e.g., principal components or admixture estimates), and do not account for local admixture patterns to inform PRS adjustment. Other limitations to clinical utility of PRS include underrepresentation of non-European ancestry individuals in GWAS, which lead to biased effect estimates when applying PRS developed in predominantly European populations to other populations, in addition to missing important population-specific variants that increase genetic risk in non-European populations^2,3,5,29,37^.

### PRIMED Consortium

The members of the Polygenic Risk Methods in Diverse Populations (PRIMED) Consortium are Clement Adebamowo, Sally Adebamowo, Adebowale Adeyemo, Nicholette Allred, Paul Auer, Jennifer Below, Palwende Romuald Boua, Kristin Boulier, Michael Bowers, Joseph Breeyear, Nilanjan Chatterjee, Brian Chen, Tinashe Chikowore, Jaewon Choi, Ananyo Choudhury, Matthew P. Conomos, David Conti, Nancy Cox, Sinead Cullina, Burcu Darst, Aaron Deutsch, Yi Ding, Todd Edwards, Eleazar Eskin, Segun Fatumo, Jose Florez, Nelson Freimer, Stephanie Fullerton, Tian Ge, Daniel Geschwind, Chris Gignoux, Stephanie Gogarten, Mark Goodarzi, Xiuqing Guo, Christopher Haiman, Neil Hanchard, Scott Hazelhurst, Ben Heavner, Susan Heckbert, Jibril Hirbo, Whitney Hornsby, Kangcheng Hou, Qinqin Huang, Alicia Huerta, Guoqian Jiang, Katherine Johnston, Linda Kachuri, Takashi Kadowaki, Abram Bunya Kamiza, Eimear Kenny, Sarah Kerns, Alyna Khan, Elena Kharitonova, Joohyun Kim, Iain Konigsberg, Charles Kooperberg, Matt Kosel, Peter Kraft, Iftikhar Kullo, Soo-Heon Kwak, Ethan Lange, Leslie Lange, Loic Le Marchand, Hyunsuk Lee, Aaron Leong, Yun Li, Meng Lin, Kirk Lohmueller, Ruth Loos, Yingchang Lu, Ravi Mandla, Alisa Manning, Iman Martin, Alicia Martin, Hilary Martin, Rasika Mathias, James Meigs, Josep Mercader, Rachel Mester, Mariah Meyer, Tyne Miller-Fleming, Braxton Mitchell, Nicola Mulder, Jie Na, Pradeep Natarajan, Sarah C. Nelson, Maggie Ng, Kristján Norland, Franklin Ockerman, Loes Olde Loohuis, Suna Onengut-Gumuscu, Ebuka Onyenobi, Roel Ophoff, Paivi Pajukanta, Bogdan Pasaniuc, Bogdan Pasaniuc, Aniruddh Patel, Ulrike Peters, Jimmy Phuong, Michael Preuss, Laura Raffield, Michèle Ramsay, Alexander Reiner, Kenneth Rice, Stephen Rich, Jerome Rotter, Bryce Rowan, Robb Rowley, Yunfeng Ruan, Lori Sakoda, Sriram Sankararaman, Dan Schaid, Dan Schrider, Philip Schroeder, Ruhollah Shemirani, Jonathan Shortt, Megan Shuey, Xueling Sim, Roelof A.J. Smit, Johanna Smith, Lucia Sobrin, Lauren Stalbow, Adrienne Stilp, Daniel Stram, Quan Sun, Ken Suzuki, Lukasz Szczerbinski, Ran Tao, Bamidele Tayo, Timothy Thornton, Buu Truong, Teresa Tusié, Miriam Udler, David van Heel, Luciana B. Vargas, Vidhya Venkateswaran, Ying Wang, Jia Wen, Jennifer Wessel, Laura Wiley, Lynne Wilkens, Riley Wilson, John Witte, Genevieve Wojcik, Quenna Wong, Toshimasa Yamauchi, Lisa Yanek, Yue Yu, Yuji Zhang, Haoyu Zhang, and Michael Zhong.

## Acknowledgements

This work utilized data generated by the Reasons for Geographic and Racial Differences in Stroke (REGARDS) project. This research project is supported by cooperative agreement U01 NS041588 co-funded by the National Institute of Neurological Disorders and Stroke (NINDS) and the National Institute on Aging (NIA), National Institutes of Health, Department of Health and Human Service. The content is solely the responsibility of the authors and does not necessarily represent the official views of the NINDS or the NIA. Representatives of the NINDS were involved in the review of the manuscript but were not directly involved in the collection, management, analysis or interpretation of the data. The authors thank the other investigators, the staff, and the participants of the REGARDS study for their valuable contributions. A full list of participating REGARDS investigators and institutions can be found at: https://www.uab.edu/soph/regardsstudy/. The REGARDS study protocol was reviewed and approved by the Institutional Review Board at the University of Alabama at Birmingham. All participants provided written informed consent, and all data used for these analyses were previously de-identified.

## Data availability

REGARDS data are not publicly available due to ethical and legal restrictions. To abide by its obligations with NIH/NINDS and the Institutional Review Board of the University of Alabama at Birmingham, REGARDS facilitates data sharing through data use agreements. Any investigator is welcome to access the REGARDS data, including statistical code, through this process. Requests for data access may be sent to the REGARDS Executive Committee at. More information can be found at https://www.uab.edu/soph/regardsstudy/.

