## Supplemental File for "Ancestry Calibration of Polygenic Risk Scores Improves Risk Stratification and Effect Estimation in African American Adults"

Luciana B. Vargas *et al.*

### Table of Contents

Extended Methods ... 2
- Study population ... 2
- Genetic analyses ... 2
- Polygenic Risk Score calculation ... 4
- Statistical Analysis ... 4

Supplementary Tables ... 6
Supplementary Figures ... 8
References ... 15

### Extended Methods

#### *Study population*

The Reasons for Geographic and Racial Differences in Stroke (REGARDS) cohort is a population-based prospective cohort designed to study disparities in the incidence of stroke and related comorbidities across the U.S.[1] Recruitment included over 30,000 individuals aged 45 years or older living in the continental U.S. and enriched for those living in the “Stroke belt” (Arkansas, Alabama, Georgia, Louisiana, Mississippi, North Carolina, South Carolina, and Tennessee)[1]. In REGARDS, participants’ phenotypes have been extensively characterized and have greater than 10 years follow-up. In this study, we included REGARDS individuals who self-identified as “Black” or African American, and had genetic data available (N = 8,843). The three outcomes, height, T2D, and fasting glucose, were analyzed cross-sectionally (baseline measure) at visit 1 and were assessed through REGARDS study interviews, in-home measurements, and laboratory testing. Diabetes was defined as fasting glucose ≥126 mg/dL or non-fasting glucose ≥ 200 mg/dL, or self-reported use of diabetes medications. Diabetes status was available for 8,684 individuals. Continuous glucose (mg/dL) was collected from blood and evaluated in fasting non-diabetic individuals (N = 5,258). For height (inches), we excluded individuals older than 90 years, given previous observations that height decreases not linearly at older ages[2,3]. Additionally, we excluded individuals with more than 4 standard deviations (s.d.) from the mean height once stratified by sex and race (N = 8,738). Social determinants of health (SDoH) data were obtained via self-reported computer-assisted telephone interviews (CATI) conducted during the second study visit. Income was classified into four categories (<$20K, $20K-$35K, $35K-75K, $75K+), education was classified into four categories (lower than high school, completed high school, some college, completed college and above), and medical care was defined as whether the individual had a clinic or doctor who provided regular medical care. The REGARDS study protocol was reviewed and approved by the Institutional Review Board at the University of Alabama at Birmingham. All participants provided written informed consent, and all data used for this analysis was previously de-identified.

#### *Genetic analyses*

Blood samples from the REGARDS baseline study visit (2003-2007) were centrifuged and shipped cold by overnight courier to the Laboratory for Clinical Biochemistry Research (LCBR) at the University of Vermont where they were logged into the database, processed, aliquoted into 0.5 mL and 1.0 mL cryovials, and stored at -80˚C. DNA was extracted from EDTA packed cells using Qiagen (Hilden, Germany) extraction kits. Genotyping was performed using the

Expanded Multi-ethnic Genotyping Array ($MEGA^{EX}$) (Illumina, San Diego, CA). The $MEGA^{EX}$ chip provides genotyping on ~1.8 million markers from various ancestry populations, making it highly suitable for diverse and admixed populations. GenomeStudio Software v2.04 (Illumina, San Diego, CA) was used for SNP calling. We cleaned genetic data in PLINK (v. 2)[4] to remove related individuals (pairwise IBD > 0.1875) and selected genetic variants (AT-CG, multiallelic variants and poorly genotyped variants [missing in >2% of individuals genotyped]). Then, genetic data on 8,843 self-identified non-Hispanic Black participants from the REGARDS cohort[1] was imputed to TOPMed version R2 on GRCh38[5,6]. Post imputation, we removed poorly imputed variants [INFO < 0.3]. The cleaned imputed genetic dataset included 75,233,979 variants.

For genetic similarity estimates (GSYRI and PCs), variants were pruned based on estimated linkage disequilibrium (LD) (removed variants with a $r^2 > 0.1$ with another included variant in a 100 base pair window) and a minor allele frequency (MAF) greater than 5%. Principal component analysis (PCA) was used to summarize genetic data using PLINK (v. 2)[4]. The estimated proportion of genetic similarity to YRI (GSYRI) was derived from maximum likelihood estimation of ancestry proportions with predefined K=2, in the software ADMIXTURE (v. 1.3)[7,8]. Based on known demographic history of African American individuals in the U.S., we assumed two independent subpopulations of contributing ancestries (K=2) and included 99 Northern European from Utah (CEU) and 108 Yoruba in Ibadan, Nigeria (YRI) genomes from 1000 Genomes panel[9] as reference populations. These populations are commonly used to estimate continent-level genetic ancestry for individuals living in the United States and are treated as proxies for ancestral populations. It is important to note these are modern-day human genomes and they do not fully capture the European and African historical genetic continental contributions to African Americans living in the U.S. Given these limitations, we describe our REGARDS participants using GSYRI as opposed to the alternative nomenclature commonly used, “estimated proportion of African ancestry”. It was verified that including additional 1,000 Genomes reference populations and expanding the number of populations (K>2) had no significant impact on the estimated GSYRI nor did we find evidence for additional significant ancestry contributions in REGARDS self-identified non-Hispanic Black study participants. We further note that our estimates of ancestry proportions using K=2 supervised clustering were highly correlated with our estimates of ancestry proportions using K=2 unsupervised clustering in ADMIXTURE, even when excluding the 1,000 Genomes reference populations. We also assessed genetic similarity by deriving principal components (PC) from REGARDS and 1,000 Genomes genetic data in PLINK (v. 2)[4].

***Polygenic Risk Score calculation***

Two recent PRS functions based on large multiethnic GWAS studies were retrieved from the PGSCatalog[10], one for height (PGS002802)[11] and one for type 2 diabetes (T2D) (PGS002308)[12]. $PRS_{height}$ was developed by applying SBayesC[13] to estimate joint SNP effects through meta-analysis which included 5,314,291 individuals of primarily European (75.8%), followed by East Asian (8.8%), Hispanic (8.5%), African (5.5%) and South Asian (1.4%) ancestry or ethnicity groups[11]. While $PRS_{T2D}$ was developed using PRS-CSx[14], a method that estimates SNP effects through Bayesian regression in different populations while implementing a shared continuous shrinkage (CS) prior, which accounts for varying patterns of linkage disequilibrium between ancestral groups[15]. $PRS_{T2D}$ meta-analysis included 1,099,372 individuals of primarily European (81.7%), African American (2.2%) and Japanese (16.1%) ancestry, ethnicity or nationality groups[12]. There were 1,254,608 variants in REGARDS that overlapped with the 1,259,754 variants in the $PRS_{T2D}$ score (99.6%). For $PRS_{height}$, 1,016,335 variants were present in REGARDS of the total 1,103,042 present in the $PRS_{height}$ score (92.1%). We applied PRS functions to REGARDS individuals using a custom R script following the equation $PRS_i = \sum \beta_j \times G_{j,i}$, where $i$ corresponds to REGARDS individual $i$; $\beta_j$ represents the effect size of a genetic variant $j$ on the outcome evaluated by the PRS; and $G_{i,j}$ represents the risk allele count for variant $j$ in individual $i$ (0, 1 or 2). For each variant, the alleles in REGARDS were flipped to match the risk allele in each score before calculation. To facilitate comparisons between calibrated and uncalibrated PRS, we standardized PRS values across all AA individuals (ignoring GSYRI decile groupings) to a mean of 0 and standard deviation of 1. We validated our calibration approach by comparing PRS residuals after regressing out GSYRI to those obtained by regressing out PC 1-10 and by using the Polygenic Score Catalog Calculator[16] ancestry normalization tool[17].

***Statistical Analysis***

We evaluated the performance of $PRS_{height}$ in relation to height (inches), and $PRS_{T2D}$ in the context of T2D status and in relation to glucose levels (mg/dL) in fasting non-diabetic individuals. For each outcome, we built a series of linear (height, glucose) and logistic (T2D) regression models, with the stepwise addition of i) basic covariates (sex and age); ii) basic covariates and GSYRI; iii) basic covariates and PRS; iv) basic covariates, GSYRI and PRS; and finally, v) basic covariates, GSYRI, PRS, and an interaction term between GSYRI and PRS. In models evaluating T2D and glucose, BMI was also included as a basic covariate. PRS were analyzed continuously, and categorized by deciles of its distribution. Regression models and Pearson's correlations were computed using the R *stats* package[18]. We computed variance explained ($R^2$), area under the receiver operator curve (AUC), odds ratio (OR), and effect estimates for the different calibrated and uncalibrated PRS to compare overall model prediction (using *rsq*[19], *epitools*[20] and *pROC*[21] packages). We also evaluated PRS performance in the context of classification bias, by comparing the observed versus expected percentage of samples classified "at-risk" by the PRS (in the top decile of the PRS distribution) by percentile ranges of GSYRI (<50%, 50-60%, 60-70%, 70-80%, 80-90%, ≧90%), where we would expect 10% of individuals to be classified "at-risk" independent of GSYRI groupings (for those outcomes not associated with GSYRI, height and glucose, or showing a different direction of effect, as in T2D). We applied difference-of-odds-ratios (DOR) test[22] to formally compare uncalibrated and calibrated PRS ability to classify cases in the upper 10% vs. lower 10%, where a *p*-value can be obtained from the *z*-score distribution by using the formula $z = (\beta_1 - \beta_2)/\sqrt{\sigma_1{}^2 + \sigma_2{}^2}$.

**Supplementary Tables**

| | **Females** | **Males** | **Overall** |
|---|---|---|---|
| Sample size (n (%)) | 5,394 (60.1%) | 3,449 (39.0%) | 8,843 |
| Age (mean ± s.d.) | 63.3 ± 9.3 | 64.0 ± 9.1 | 63.6 ± 9.2 |
| Genetic similarity to YRI, GSYRI (median [IQR]) | 84.5 [75.7 - 90.7] | 84.0 [75.1-90.4] | 84.3 [75.4-90.6] |
| Height, in inches (mean ± s.d.) | 64.3 ± 2.7 | 69.8 ± 2.9 | 66.5 ± 3.8 |
| Type 2 diabetes (n (%)) | 1,517 (28.7%) | 1,047 (30.9%) | 2,564 (29.5%) |
| Fasting glucose, non-diabetics, mg/$\mu$L (mean ± s.d.) | 93.2 ± 11.2 | 93.7 ± 11.5 | 93.4 ± 11.3 |

**Supplementary Table 1.** Demographic and clinical characteristics of self-identified African American individuals from the REasons for Geographical and Racial Disparities in Stroke (REGARDS) cohort, stratified by biological sex. YRI, Yoruba in Ibadan, Nigeria, 1000 Genomes Project. s.d., standard deviation. IQR, interquartile range.

| | **< 50%** | **50-60%** | **60-70%** | **70-80%** | **80-90%** | **> 90%** | **All** |
|---|---|---|---|---|---|---|---|
| **Sample size** | 244 | 323 | 781 | 1,754 | 3,220 | 2,410 | 8,732 |
| **PRS$_{T2D}$** | | | | | | | |
| Median % uncalibrated PRS | 4.9 | 11.1 | 20.3 | 35.7 | 56.0 | 69.9 | 50.0 |
| Median % calibrated PRS | 54.1 | 49.8 | 47.9 | 49.7 | 51.1 | 48.3 | 50.0 |
| **Median of the % changes** | **47.4** | **34.3** | **24.1** | **8.8** | **-4.1** | **-17.5** | **-1.8** |
| **PRS$_{height}$** | | | | | | | |
| Median % uncalibrated PRS | 95.9 | 90.1 | 81.0 | 66.1 | 45.8 | 28.0 | 50.0 |
| Median % calibrated PRS | 50.8 | 52.6 | 51.3 | 50.7 | 49.0 | 50.5 | 50.0 |
| **Median of the % changes** | **-44.0** | **-33.3** | **-24.4** | **-10.3** | **4.1** | **20.3** | **1.58** |

**Supplementary Table 2.** Percentile-based (%) ranking of individuals before and after PRS calibration. The columns represent GSYRI intervals (0-100%) and the whole cohort (All).

| | GSYRI | | PRS | | $VIF_{PRS}$ | $R^2$ (%) |
|---|---|---|---|---|---|---|
| | $\beta$ ± s.e. | *p*-value | $\beta$ ± s.e. | *p*-value | | |
| **Glucose** | | | | | | |
| ~ Age + Sex + BMI | - | - | - | - | - | 3.18 |
| + GSYRI | 0.041 ± 0.21 | 0.97 | - | - | - | 3.18 |
| + **Uncalibrated PRS** | - | - | 0.99 ± 0.16 | 2.16 x $10^{-09}$ | 1.00 | 3.85 |
| + GSYRI + **Uncalibrated PRS** | **-6.48 ± 1.49** | **1.39 x $10^{-05}$** | **1.51 ± 0.20** | **1.40 x $10^{-13}$** | **1.54** | 4.20 |
| + GSYRI + Uncalibrated PRS + **(GSYRI x Uncalibrated PRS)** | -6.26 ± 1.71 | 2.60 x $10^{-04}$ | 1.29 ± 0.88 | 0.14 | - | 4.20 |
| | Interaction: $\beta$ ± s.e. = 0.28 ± 1.07, *p* = 0.79 | | | | | |
| + **Calibrated PRS** | - | - | 1.20 ± 0.16 | 2.12 x $10^{-13}$ | 1.00 | 4.18 |
| + GSYRI + **Calibrated PRS** | 0.36 ± 1.20 | 0.77 | 1.20 ± 0.16 | 2.03 x $10^{-13}$ | 1.00 | 4.19 |
| + GSYRI + Calibrated PRS + **(GSYRI x Calibrated PRS)** | 0.16 ± 1.22 | 0.90 | 2.36 ± 1.06 | 0.026 | - | 4.21 |
| | Interaction: $\beta$ ± s.e. = -1.43 ± 1.29, *p* = 0.27 | | | | | |

**Supplementary Table 3.** Change in calibrated and uncalibrated $PRS_{T2D}$ model parameters with glucose as the outcome. The genetic ancestry, as represented by estimated proportion of genetic similarity to the 1,000 Genomes Yoruba panel (GSYRI), and both calibrated and uncalibrated $PRS_{T2D}$ values were included in the models one at a time, and together. $\beta$, effect size of one unit increase in the outcome. s.e., standard error. $VIF_{PRS}$, variance inflation factor for the PRS in the model. $R^2$, percentage of glucose variance explained by the model. For interaction models (noted by the inclusion of the interaction term GSYRI x PRS), parameter estimates are given for each of the main effects (GSYRI or PRS) and the interaction effect (GSYRI x PRS).

**Supplementary Figures**

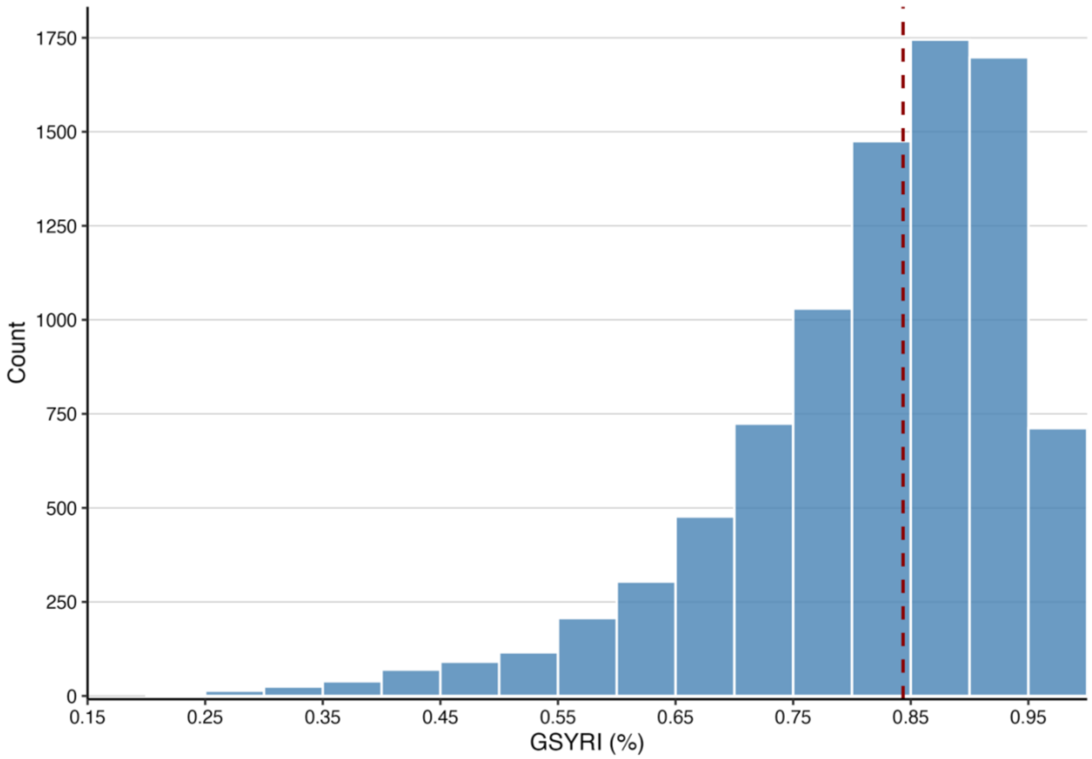


**Supplementary Figure 1.** Distribution of estimated proportion of genetic similarity to the 1,000 Genomes Yoruba panel (GSYRI) in black individuals from REGARDS. The dark red dashed line represents the median GSYRI (84.35%) in the group.

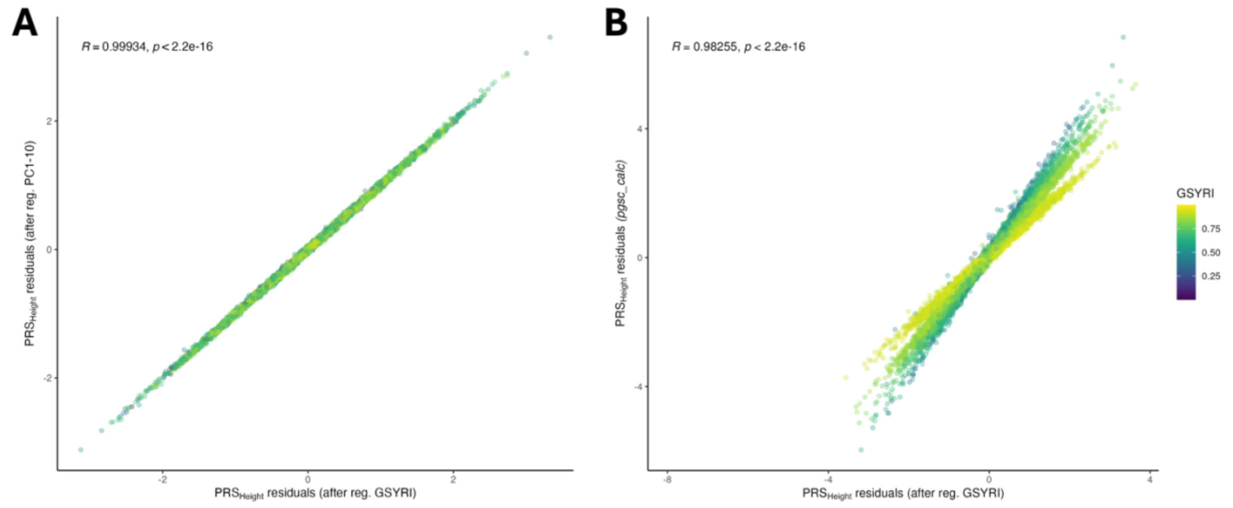


**Supplementary Figure 2**. Pearson's correlation between the $PRS_{height}$ residuals after regressing out GSYRI (*x* axis) and **A)** the 10 first principal components (PC1-10) of genetic variability (*y* axis) or **B)** using the using the Polygenic Score Catalog Calculator[16] ancestry normalization tool (*pgsc_calc*)[17]. Each point represents an individual, and the points are colored by individual's respective GSYRI (scale shown on the right). Person correlation value (R) and significance of the correlation (*p*-value) are shown at the top left corner of each plot.

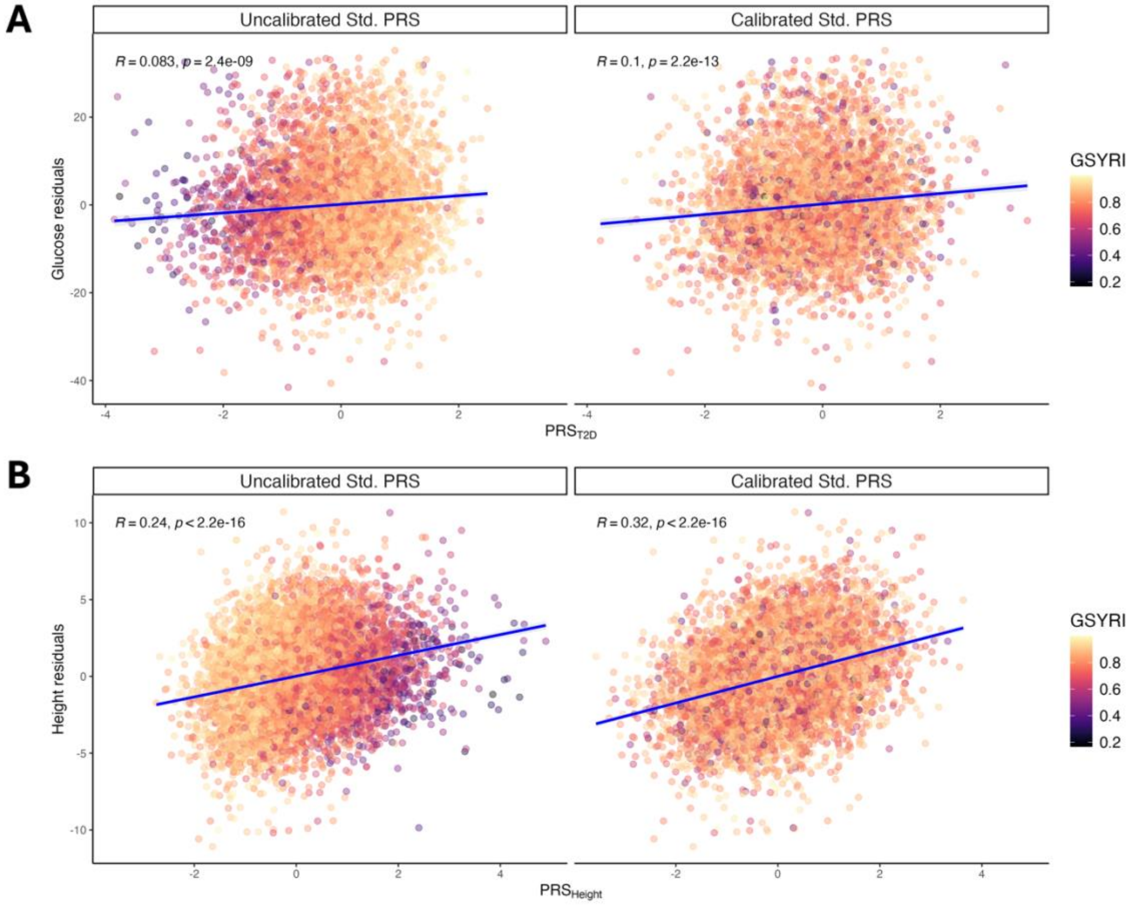


**Supplementary Figure 3.** After ancestry-adjustment (using GSYRI), PRS functions have higher correlation with residuals of **A)** glucose (mg/dL) in non-diabetic individuals and **B)** height, after regressing out sex and age. Person correlation values (R) and significance of the correlation (p-values) are shown at the top left corner.

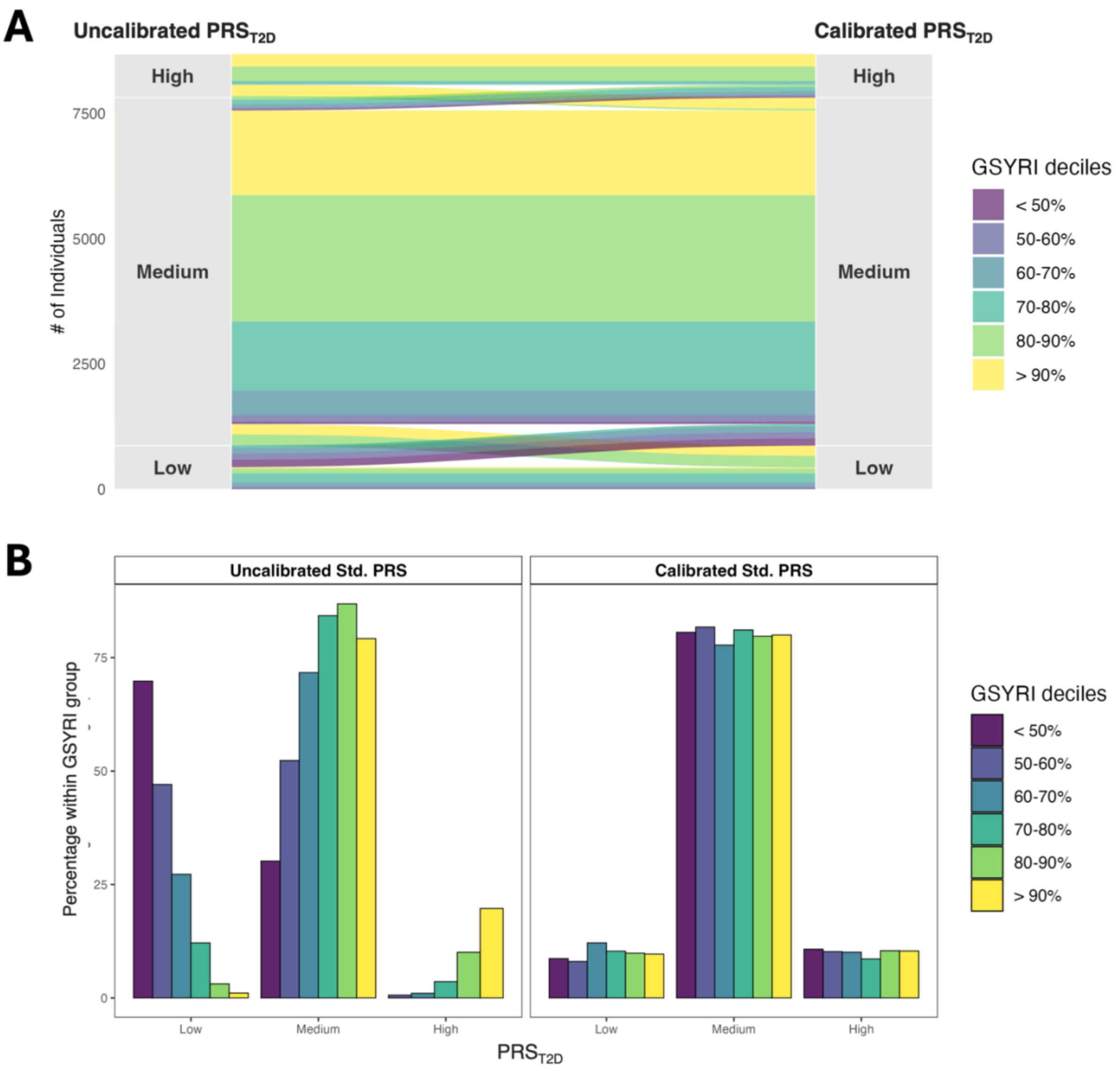


**Supplementary Figure 4. Plots showing the reclassification of individuals across $PRS_{T2D}$ risk strata (Low, Medium and High) before and after score calibration, further stratified by GSYRI (colors).** Risk categories are defined using cutoffs of the PRS distribution in the full sample: "High" is defined as the top 10%, "Medium" comprises those in 10-90% percentiles, and "Low" represents the bottom 10%. The "High" $PRS_{T2D}$ classification represents individuals at a greater risk of having type 2 diabetes. **A)** Sankey plot showing the trajectory of individuals assigned to each $PRS_{T2D}$ tertile (low, medium and high) before and after calibration. The left stratum represents ranking based on the uncalibrated $PRS_{T2D}$, and the right stratum represents ranking based on the calibrated $PRS_{T2D}$. Flows are color-coded by GSYRI groups. The width of each flow is proportional to the number of individuals transitioning between $PRS_{T2D}$ categories. **B)** Bar plot displaying the percentage of individuals within each GSYRI grouping (> 90%, 80-

90%, 70-80%, 60-70%, 50-60%, and < 50%) assigned to each $PRS_{T2D}$ tertile on the *y* axis. Within each panel, the plot on the left represents ranking based on the uncalibrated $PRS_{T2D}$, and the plot on the right represents ranking based on the calibrated $PRS_{T2D}$.

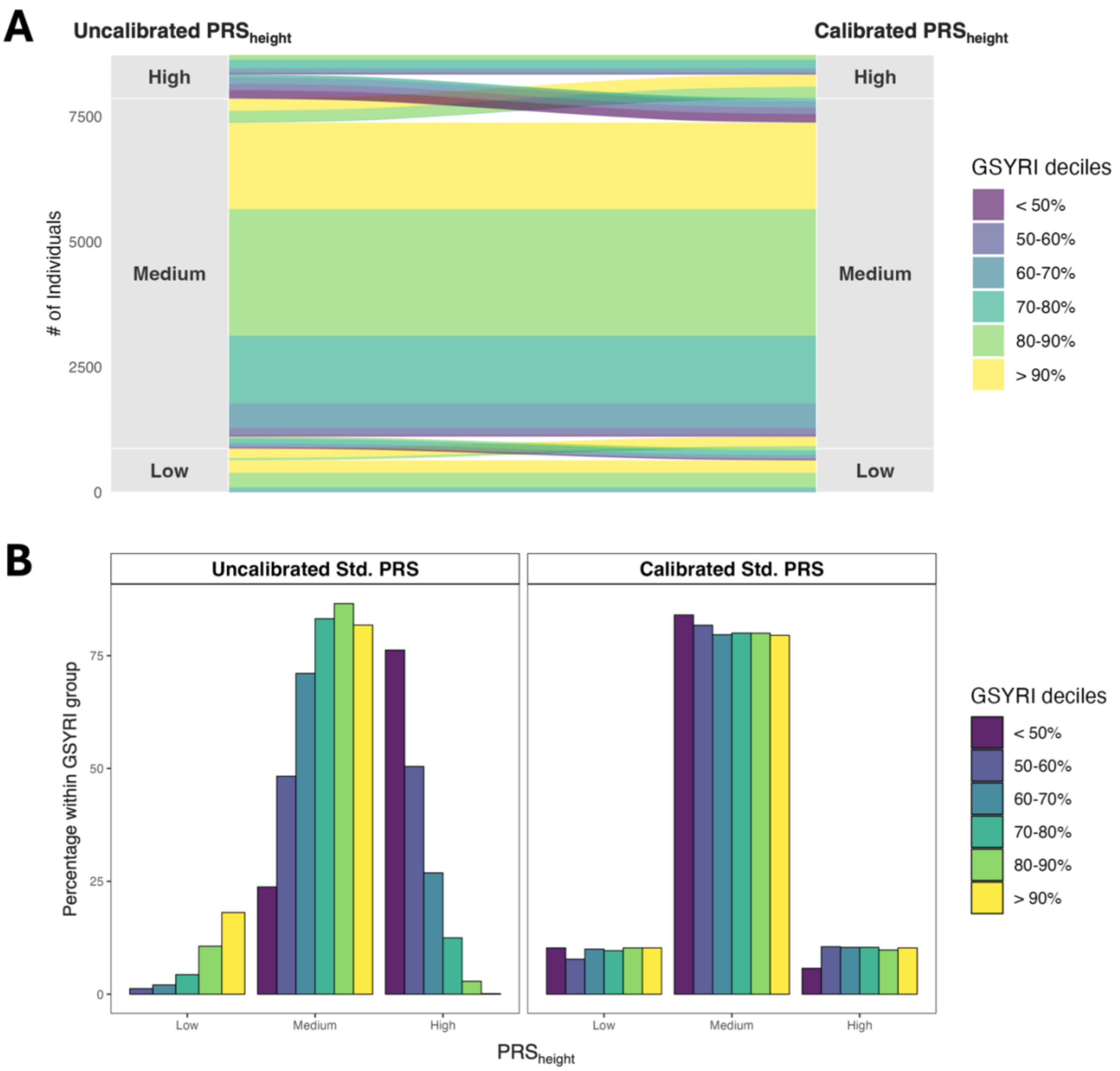


**Supplementary Figure 5. Plots showing the reclassification of individuals across $PRS_{height}$ risk strata (Low, Medium and High) before and after score calibration, further stratified by GSYRI (colors).** Risk categories are defined using cutoffs of the PRS distribution in the full sample: "High" is defined as the top 10%, "Medium" comprises those in 10-90% percentiles, and "Low" represents the bottom 10%. The "High" $PRS_{height}$ classification for height represents individuals more likely to be taller. **A)** Sankey plot showing the trajectory of individuals assigned to each $PRS_{height}$ risk stratum (low, medium and high) before and after calibration. The left stratum represents ranking based on the uncalibrated $PRS_{height}$, and the right stratum represents ranking based on the calibrated $PRS_{height}$. Flows are color-coded by GSYRI groups. The width of each flow is proportional to the number of individuals transitioning between $PRS_{height}$ categories. **B)** Bar plot displaying the percentage of individuals within each GSYRI grouping (> 90%, 80-

90%, 70-80%, 60-70%, 50-60%, and < 50%) assigned to each $PRS_{height}$ risk stratum on the *y* axis. Within each panel, the plot on the left represents ranking based on the uncalibrated $PRS_{height}$, and the plot on the right represents ranking based on the calibrated $PRS_{height}$.
